# Early EEG biomarkers of perinatal arterial ischemic stroke:a case series

**DOI:** 10.1101/2023.03.24.23287727

**Authors:** Xiu-Ying Fang, Qiong Wu, Ying-Jie Wang, Jian Mao

**Affiliations:** Department of Neurology, Shengjing Hospital of China Medical University, Shenyang, China; Department of Neonatal Pediatrics, Shengjing Hospital of China Medical University, Shenyang, China

**Keywords:** perinatal arterial ischemic stroke, electroencephalogram, neonate, seizure

## Abstract

**Background:** Continuous electroencephalogram (EEG) monitoring after seizures may provide an early clue to the diagnosis of perinatal arterial ischemic stroke (PAIS). We aimed to explore EEG biomarkers and seizure patterns of PAIS within 7 days.

**Methods:** Data of children with PAIS were retrospectively analyzed. Using multi-channel video electroencephalogram (vEEG) and amplitude-integrated electroencephalogram (aEEG), the patients were divided into three groups according to the monitoring start time after symptom onset: ≤48 h, 48–72 h, and 4–7 days. The characteristics of EEG activity and the law of seizures were compared across groups. Together with magnetic resonance imaging (MRI), the spatial correspondence between brain electrical activity and injury site was determined.

**Results:** Of 21 children studied, 19 (90.47%) developed disease within 48 h of delivery. Abnormal sleep–wake cycle, abnormal fast-wave activity, abnormal wave activity, hemispheric asymmetry, electrographic seizures (ESz), and electroclinical seizures (ECSz) were more common in the ≤48 h group and 48–72 h group. Differences among all groups were statistically significant (*P* <0.01). The most common abnormal wave activity was sharp wave or freak wave, consistent with the MRI-confirmed injury site. ESz and ECSz were detected in 12 cases (92.31%) in the ≤48 h group and two cases (15.38%) in the 48–72 h group, 66.67% were clonic, and 91.67% were frequent seizures. ESz or ECSz were not detected in the 4–7-day group. The starting point of ESz or ECSz was consistent with the area of the lesion shown by MRI.

**Conclusions:** Bi-hemispheric diffuse abnormal fast wave activity, sharp wave or freak wave activity in the local brain region, and ESz or ECSz initiated in the same brain region are EEG biomarkers indicating PAIS. The duration of ESz or ECSz triggered by PAIS was less than 72 h.

## 1. Introduction

Perinatal arterial ischemic stroke (PAIS) is a focal brain injury that occurs in full-term newborns and is a common cause of neonatal seizures, with an incidence of 5–18 cases per 100,000 newborns^[1-3]^. Accurate diagnosis early in the disease is the first step toward finding the appropriate treatment. Brain magnetic resonance imaging (MRI) is the gold standard for the diagnosis of PAIS, but it is often insufficient and administered after a relative delay.

Continuous electroencephalogram (EEG) monitoring after seizures may provide an early clue to the diagnosis of brain injury. The application of amplitude-integrated electroencephalography (aEEG) monitoring in neonatal neurological diseases has been studied for decades, and it can provide an important basis for the early diagnosis of brain injury and guide treatment decisions for seizure disorders. In recent years, a few studies have preliminarily described the brain electrical activity in PAIS^[3-5]^. However, due to the lack of analysis of the original electroencephalogram (EEG) waveform by aEEG, the guiding role of aEEG in the diagnosis and treatment of PAIS is limited. In addition, the low incidence of PAIS, few EEG monitoring opportunities, and different monitoring conditions all limit the application of EEG monitoring for the early diagnosis of PAIS and seizure management.

To better understand the dynamic changes in early EEG and seizure regularity after PAIS, in this study, multi-channel video electroencephalography (vEEG) and aEEG, interpreted according to the American Clinical Neurophysiology Society (ACNS) standardized neonatal EEG guidelines, were combined with brain MRI obtained 0–7 days after onset of PAIS. This allowed for visual analysis of the ictal and interictal EEG activity corresponding to the region of identified injury. To summarize, we analyzed the EEG biomarkers and seizure characteristics of neonates with PAIS in order to provide a reliable empirical basis for its diagnosis and treatment.

## 2. Methods

This study was approved by the Research Ethics Committee of Shengjing Hospital of China Medical University. Parental consent was obtained for the use of pediatric medical data in the preparation of this report. Results are reported in adherence to the Case Series guidelines for this studies^[6]^; a completed CARE Statement is included in the Supplemental Material. The data that support the findings of this study are available from the corresponding author upon reasonable request. The corresponding author has full access to the data in this study and attests to the quality and integrity of the data set and associated analyses.

### 2.1 Research participants

Data of patients with PAIS who were hospitalized in the Shengjing Hospital of China Medical University from 2018 to 2022 were retrospectively collected. The diagnosis of PAIS was confirmed by brain MRI. Exclusion criteria included: (1) cerebral infarction and watershed infarction caused by extensive vascular circulation disorder complicated with hypoxic ischemic encephalopathy, (2) periventricular leukomalacia and vascular infarction caused by intraventricular hemorrhage in premature infants, (3) lack of definite onset time or clinical symptoms, (4) no EEG monitoring performed within 7 days after onset, and (5) no brain MRI examination performed within 7 days after onset.

### 2.2 Research methods

#### 2.2.1 Clinical data collection

The medical history of all children was collected by two experienced neonatologists. Information including the clinical characteristics, pregnancy history, birth history, results of physical and auxiliary examinations, and follow-up data was collected.

#### 2.2.2 EEG monitoring and data analysis

All children completed vEEG and synchronous aEEG monitoring at least once during the 7 days of hospitalization. A Nicolet brain function monitor was used for recording, and each monitoring session was ≥4 h, which produced less interference and a good monitoring record. Electrodes were placed with reference to the standard international 10/20 system in accordance with neonatal EEG monitoring requirements, including electrode positions FP1, FP2, C3, C4, P3, P4, O1, O2, T3, T4, Cz, and Pz, while also placing electrocardiogram and upper limb electromyography electrodes. The aEEG montage comprised positions C3-O1 and C4-O2.

The patients were divided into three groups according to the length of time in between the first symptom and first EEG monitoring: ≤48 h, 48–72 h, and 4–7 days. Children in each group were monitored multiple times, and the earliest monitoring record per child was selected for analysis. The EEG records of each newborn were independently reviewed by experienced EEG physicians. According to the guidelines for continuous EEG monitoring of newborns issued by the ACNS^[7]^, changes in all aspects of EEG activity were analyzed, including sleep–wake cycling (SWC), voltage, physiological and abnormal wave activity, continuity, interburst interval (IBI) duration, symmetry, synchronization, and seizure type. According to the International League Against Epilepsy classification criteria for neonatal seizures^[8]^, seizures were classified as electroclinical seizure (ECSz) and electrographic seizure (ESz). ECSz is defined as a clinical seizure accompanied by abnormal EEG evolution. ESz is defined as a clear evolution of aberrant EEG activity but with no corresponding motor manifestations. Each 4-h monitoring period was rated according to the number of seizures: “occasional” (≤2 times), “multiple” (3–6 times), or “frequent” (≥7 times). Status epilepticus (SE) was determined to be present when the summed duration of seizures comprised ≥50% of an arbitrarily-defined 1-h epoch^[7]^.

Brief rhythmic discharges (BRDs) were defined as very brief (<10 s) runs of focal or generalized sharply-contoured rhythmic activity, with or without evolution, that were not consistent with any known normal or benign pattern, which have a frequency >4 Hz in adults^[9]^. Periodic discharges (PDs) were defined as patterns with waveforms of relatively uniform morphology and duration, repeating at nearly regular, quantifiable intervals^[7]^.

#### 2.2.3 Brain MRI examination

MRI was performed using a Philips Intera Achieva 3.0 T scanner, and conventional T1-weighted, T2-weighted, and diffusion-weighted imaging sequences were collected. All MRI findings were analyzed by neonatal neurologists and imaging experts. Injury sites were classified according to their anatomical location^[10]^.

### 2.3 Statistical analysis

SPSS 22.0 software was used for statistical analysis. Normally-distributed data are expressed as means ± standard deviations, whereas non-normally-distributed data are expressed as medians and interquartile ranges. Cochran’s Q test was used for comparisons among multiple groups. *P* <0.05 was considered statistically significant.

## 3. Results

Clinical information was collected from 33 children. Of these cases,12 patients were excluded (3 children with vascular infarction caused by brain parenchyma or intraventricular hemorrhage, 5 children without clear onset time, and 4 children without EEG monitoring within 7 days after onset),leaving 21 eligible patients in the sample. There were 13 males (61.9%) and eight females (38.1%), with an average weight of 3,771 ± 571 g and gestational age of 38.9 ± 1.5 weeks. The first symptoms appeared within 24 h after birth in 11 cases (52.38%), 24–48 h after birth in eight cases (38.09%), 3–4 days after birth in one case (4.76 %), and 5 days after birth in one case (4.76%). The initial clinical symptoms were unilateral or bilateral limb shaking in 18 cases (85.71%), cyanosis of the face or acrocyanosis in three cases (14.29%), apnea in one case (4.76%), limb rigidity in one case (4.76%), and jaundice in one case (4.76%). There was no obvious history of abnormal pregnancy in any of the 21 cases. The possible factors related to PAIS after birth included ABO hemolysis in two cases (9.52%), macrosomia in one case (4.76%), subarachnoid hemorrhage in one case (4.76%), and hyperbilirubinemia in one case (4.76%).

### 3.1 EEG background activities in different periods

Three groups of neonates that underwent vEEG and aEEG monitoring at different time periods (≤48 h, 48–72 h, and 4–7 days after onset) were selected for analysis, with 13 EEG data points in each group. There were significant differences in the proportion of abnormal SWC, abnormal active fast-wave activity, abnormal symmetry, and ESz or ECSz among the three groups (*P* <0.01). The proportion of the above abnormalities was highest in the ≤48-h group, followed by the 48–72-h group.

In the ≤48-h and 48–72-h groups, the abnormal wave activity was relatively obvious, and the high-amplitude sharp wave or freak wave was the most common. BRD occurred in three cases (23.08%), and PDs occurred in two cases (15.38%). In the 4–7-day group, the abnormal wave activity decreased or disappeared, and low-amplitude irregular wave activity was dominant.

The abnormal rates of voltage, synchronization, continuity, and IBI were relatively low in the whole sample, and there were no significant differences among the three groups (*P* >0.05). The EEG background activity in each group is presented in Table 1.

**Table 1.**
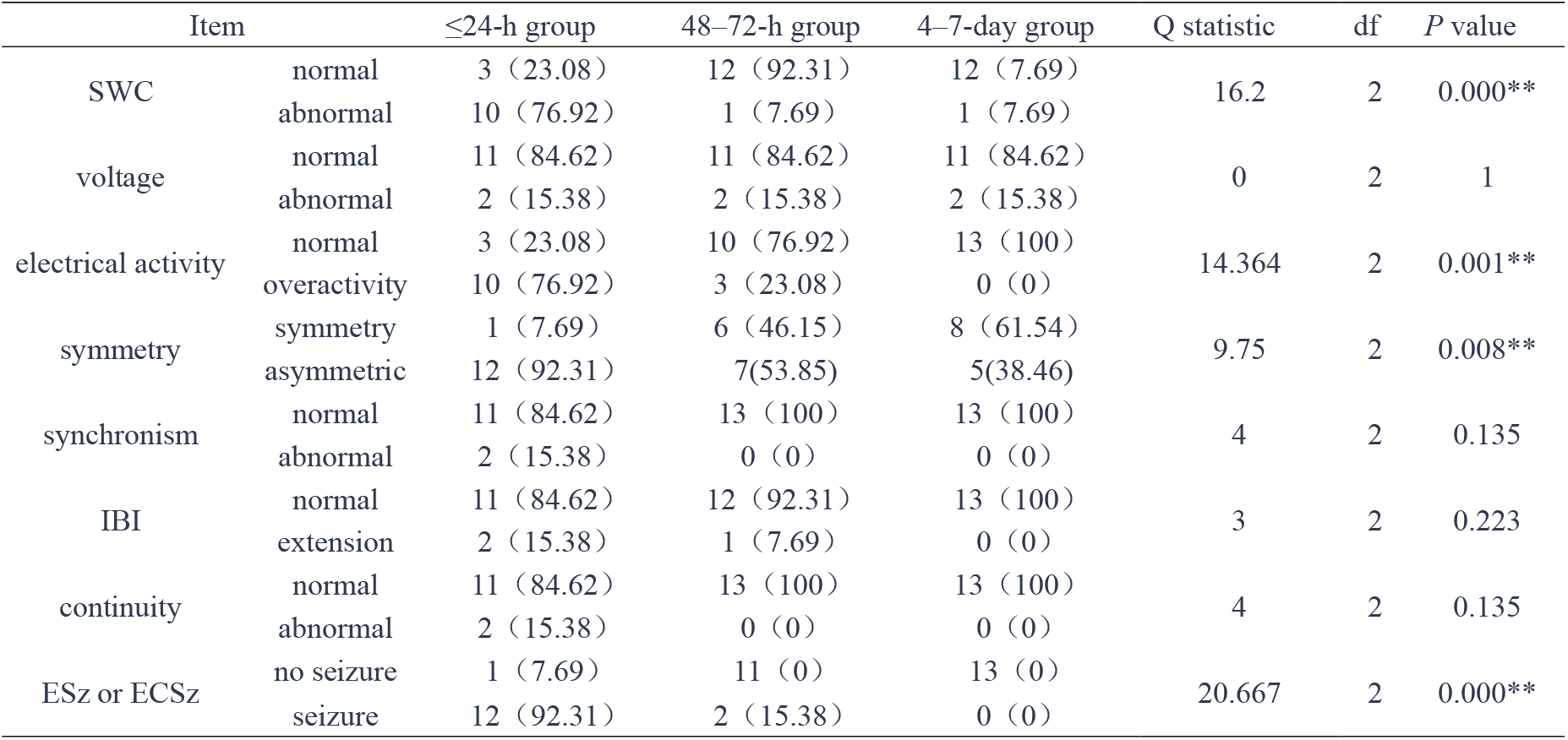
Comparison of electroencephalogram (EEG) monitoring background activity. SWC, sleep-wake cycle; IBI, interburst interval; Esz, electrographic seizures; ECSz, electroclinical seizures

| Item |  | ≤24-h group | 48–72-h group | 4–7-day group | Q statistic | df | P value |
| --- | --- | --- | --- | --- | --- | --- | --- |
| SWC | normal | 3 (23.08) | 12 (92.31) | 12 (7.69) | 16.2 | 2 | 0.000** |
|  | abnormal | 10 (76.92) | 1 (7.69) | 1 (7.69) |  |  |  |
| voltage | normal | 11 (84.62) | 11 (84.62) | 11 (84.62) | 0 | 2 | 1 |
|  | abnormal | 2 (15.38) | 2 (15.38) | 2 (15.38) |  |  |  |
| electrical activity | normal | 3 (23.08) | 10 (76.92) | 13 (100) | 14.364 | 2 | 0.001** |
|  | overactivity | 10 (76.92) | 3 (23.08) | 0 (0) |  |  |  |
| symmetry | symmetry | 1 (7.69) | 6 (46.15) | 8 (61.54) | 9.75 | 2 | 0.008** |
|  | asymmetric | 12 (92.31) | 7(53.85) | 5(38.46) |  |  |  |
| synchronism | normal | 11 (84.62) | 13 (100) | 13 (100) | 4 | 2 | 0.135 |
|  | abnormal | 2 (15.38) | 0 (0) | 0 (0) |  |  |  |
| IBI | normal | 11 (84.62) | 12 (92.31) | 13 (100) | 3 | 2 | 0.223 |
|  | extension | 2 (15.38) | 1 (7.69) | 0 (0) |  |  |  |
| continuity | normal | 11 (84.62) | 13 (100) | 13 (100) | 4 | 2 | 0.135 |
|  | abnormal | 2 (15.38) | 0 (0) | 0 (0) |  |  |  |
| ESz or ECSz | no seizure | 1 (7.69) | 11 (0) | 13 (0) | 20.667 | 2 | 0.000** |
|  | seizure | 12 (92.31) | 2 (15.38) | 0 (0) |  |  |  |

### 3.2 ESz and ECSz

In the ≤48-h group, 92.31% (12/13) experienced ESz or ECSz during monitoring. There were 16.67% (2/12) of only ESz, 58.33% (7/12) of only ECSz, and 25% (3/12) of ESz and ECSz. ECSz included clonic seizures (8/12, 66.67%), automatic seizures (2/12, 16.67%), and sequential seizures (1/12, 8.33%). Of the recorded seizures, 91.67% (11/12) were frequent seizures, and 8.33% (1/12) were multiple seizures, none of which reached SE. In the 48–72-h group, 15.38% (2/13) had occasional seizures; the interval between seizures was prolonged, the amplitude of seizures decreased, and all of them were transient ESz. No ESz or ECSz were detected in the 4–7-day group.

### 3.3 Brain MRI and EEG

For each patient, the first brain MRI examination was completed within 0–7 days (2.3 ± 2.1 days) after onset. There were 15 cases (71.43%) of left-sided stroke, five cases (23.81%) of right-sided stroke, and one case (4.76%) involving bilateral lesions. In three cases (14.29%), the lesions spanned most of the unilateral cerebral hemisphere, in four cases (19.05%) the lesion was situated before the central sulcus, in five cases (23.81%) the lesion was situated after the central sulcus, in seven cases (33.33%) the lesions were clustered anterior and posterior to the central sulcus, and in one case (4.76%) the lesion was located in the occipital region.

The initial side of electrical evolution in the 12 cases of ESz or ECSz was consistent with the injured side as shown by brain MRI. Abnormal waves were observed in 18 cases (85.71%), and were consistent with the location of the injury. No obvious abnormal wave activity was observed in three cases (14.29%). A comparison of abnormal wave activity sites and seizures corresponding with brain MRI results is shown in Table 2.

**Table 2.**
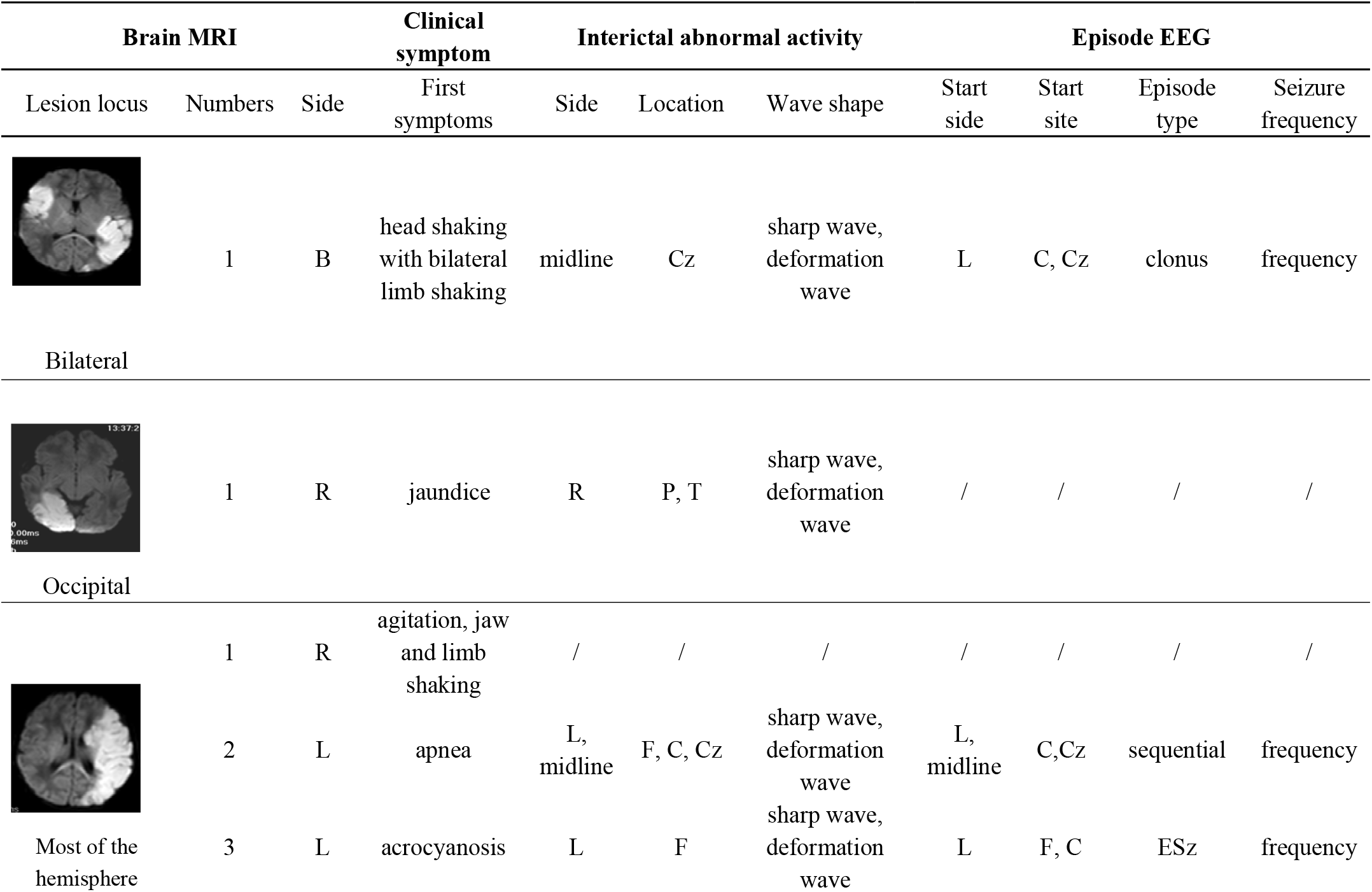
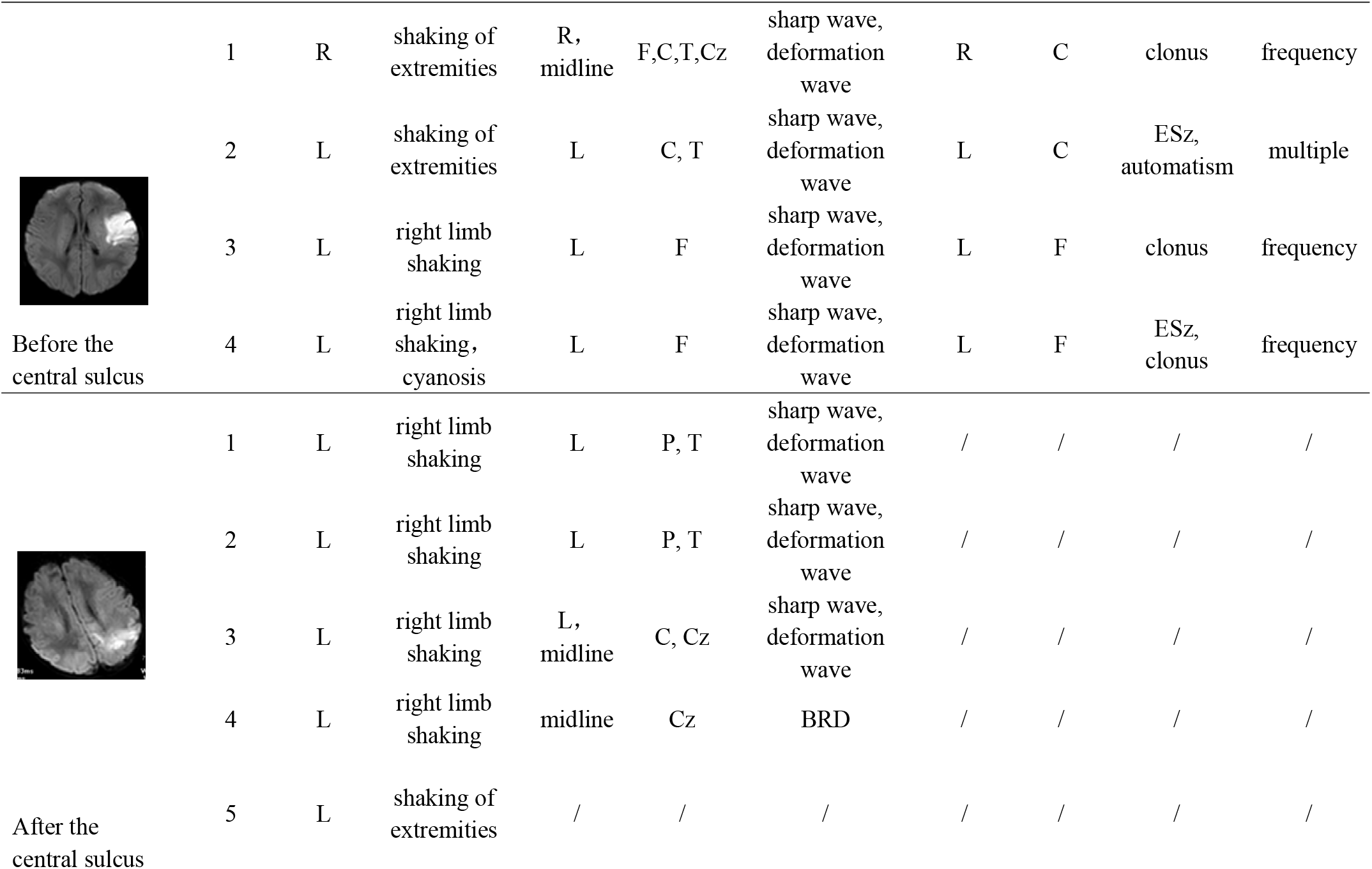
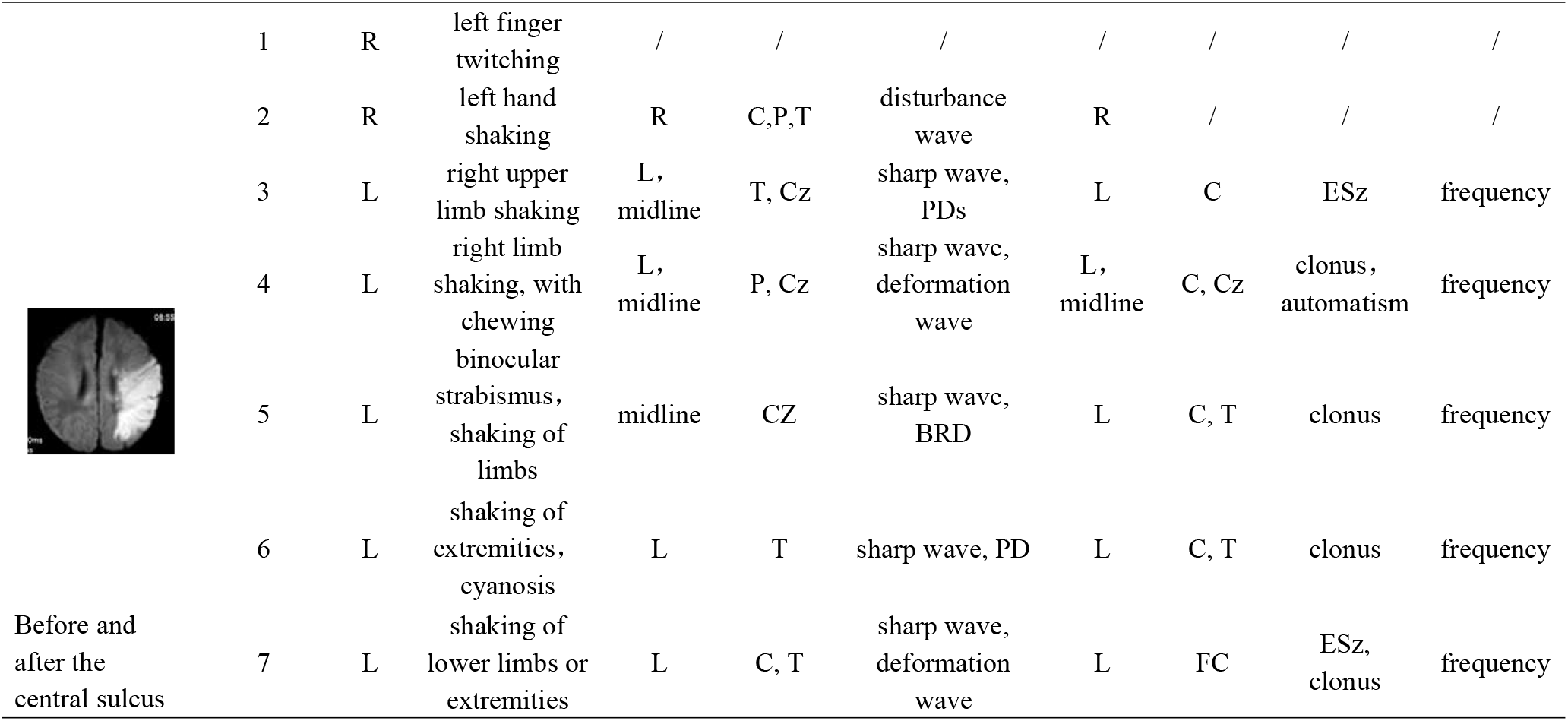
Stroke site versus interictal abnormal wave activity and seizure. MRI, magnetic resonance imaging; EEG, electroencephalogram; M, midline; L, left; R, right; C, central; F, frontal; P, parietal; O, occipital; T, temporal; Cz, central zone; Esz, electrographic seizures; PDs, periodic discharges; BRD, brief rhythmic discharge

### 3.4 Follow-up

Four children in the sample were reexamined with vEEG, aEEG, and brain MRI 1–4 months after birth. MRI showed extensive softening at the previous stroke site. Physical examination showed that the muscle strength of the contralateral limb was weaker than that of the healthy side, the tendon reflex was slightly reduced, and activity was relatively reduced. The assessment of mental intelligence development was almost normal. The vEEG and aEEG examination showed a significant reduction in EEG activity in the lesion area and the disappearance of indicative physiological wave activity. No ESz or ECSz were detected in the four patients.

## 4. Discussion

PAIS is a common cause of neonatal seizures. EEG monitoring provides important information for assessment of neonatal cerebral function and injury. In this study, continuous dynamic monitoring using multi-channel vEEG and aEEG revealed that, over the course of 7 days, EEG background activity and seizures changed significantly. During the acute stage, diffuse hyperactive background activity in both hemispheres, high-amplitude sharp or freak wave activity in local brain regions, and ESz or ECSz initiated in the same region often indicated the presence of PAIS and the injured side.

Advances in modern neuroimaging have significantly improved the detection and recognition of PAIS. An international child stroke study found that neonatal cerebral infarction preferentially involved the middle cerebral artery and left hemisphere, with bilateral lesions accounting for only 24% of cases^[11]^. In this study, 16 cases (76.19%) involved the left middle cerebral artery, only one case (6.67%) involved the posterior cerebral artery, and no cases involved the anterior cerebral artery, similar to previous studies^[12-13]^.

Continuous dynamic EEG monitoring can reflect the severity of and recovery from brain injury in real time. The metabolism, inflammation, cell death, and other aspects caused by local hypoxia and ischemia in the brain undergo dramatic dynamic changes over a short period of time. Our study found that the effect of PAIS on brain function in the acute phase is not limited to the injured side, and the contralateral hemisphere also changes accordingly. Within 48 h of onset, the brain electrical activity was significantly abnormal, and the asymmetry of amplitude or waveform between the two hemispheres was most obvious. During this period, β wave activity increased significantly, most prominently in the affected hemisphere. José et al. confirmed by EEG spectrum analysis that there was a significant difference in the β-band between the ischemic and unaffected sides^[3]^. Through continuous monitoring, it was found that the voltage, continuity, and IBI changed slightly at different stages after symptom onset, which has limited diagnostic value for stroke.

Local sharp waves, malformation waves, BRD, and PDs were the most common forms of abnormal EEG activity, typically appearing within 72 h of onset. Some studies have suggested that excessive focal θ activity, phase sharp wave in the Rolandic region, and PDs may be signature EEG features of PAIS^[4-5,14]^. However, through continuous dynamic observation, this study found that abnormal EEG activity is closely correlated with the anatomical location of injury and the timing of monitoring after brain injury. Through continuous EEG monitoring, it was found that these abnormal EEG activities are significantly reduced or disappear in the course of 4–7 days, with changed morphology. In addition, this study found that relatively regular discharges, such as BRD and PDs, also occurred in areas adjacent to the injury site. Consistent with this, animal models have shown that regular electrical activity such as PDs are produced in the penumbra of the injury site and are homologous to the onset of seizures^[15]^. Therefore, when multiple forms of abnormal wave activity and seizure origin are concentrated in the same area, it is highly suggestive of local cerebral cortex injury.

Neonatal seizures are the result of potential brain injury and may be the only symptom for most children with PAIS. Long-term vEEG monitoring has shown that limb clonic seizure was the most common seizure type and the most easily recognized seizure presentation. In addition, non-specific movements, such as apnea, cyanosis of limbs or faces, and chewing, can be the only symptoms to accompany the seizure. In the past, the judgment of neonatal seizures and seizure control was mainly based on the observation of neonatal behavior^[11]^, which was not confirmed by EEG monitoring. Therefore, there is no clear consensus as to when seizures completely disappear in children with PAIS and when to stop the use of anticonvulsant drugs. In this study, vEEG monitoring found that after 48 h of onset, seizures gradually abated. Neither ESz nor ECSz were detected 4–7 days after onset. It is speculated that ESz or ECSz caused by PAIS may last <72 h. Therefore, for newborns with suspected PAIS, vEEG and aEEG monitoring should be implemented as soon as possible, and refined management of anticonvulsants should be achieved under vEEG monitoring. This can reduce excessive exposure to anticonvulsants and provide a reliable reference for the rational allocation of EEG monitoring resources.

Long-term follow-up of children with PAIS has revealed that 16.4% of children later developed symptomatic epilepsy. The median onset of epilepsy is 2 years after stroke^[16]^, and the risk of epilepsy is highest after 6 months^[17]^. Given this, four children in our sample were reexamined by EEG at 1–4 months of age. No ESz or ECSz were observed, which may be related to the relatively early re-examination time. Whether secondary epilepsy will occur in this group of children in the future requires close long-term follow-up.

This study has some limitations. Owing to inconsistent monitoring opportunities and timing in retrospective studies, most children in the sample lacked complete and consistent EEG monitoring data, and, because of the limited number of cases, we could not link the location or size of the infarction with the results. Whether these features have different effects on electrical activity in the brain cannot be reliably determined. Whether EEG monitoring during different periods of chronic recovery has prognostic value remains to be further studied.

To our knowledge, this is the first study to systematically summarize the chronological characteristics of EEG background activity and seizures within 0–7 days after PAIS onset. This study showed that vEEG combined with aEEG monitoring technology has broad application for the diagnosis of acute PAIS and can enhance our understanding of disease mechanisms. EEG monitoring provides a reliable diagnostic basis, especially for individualized, time-sensitive management of PAIS seizures.

**Figure 1.**
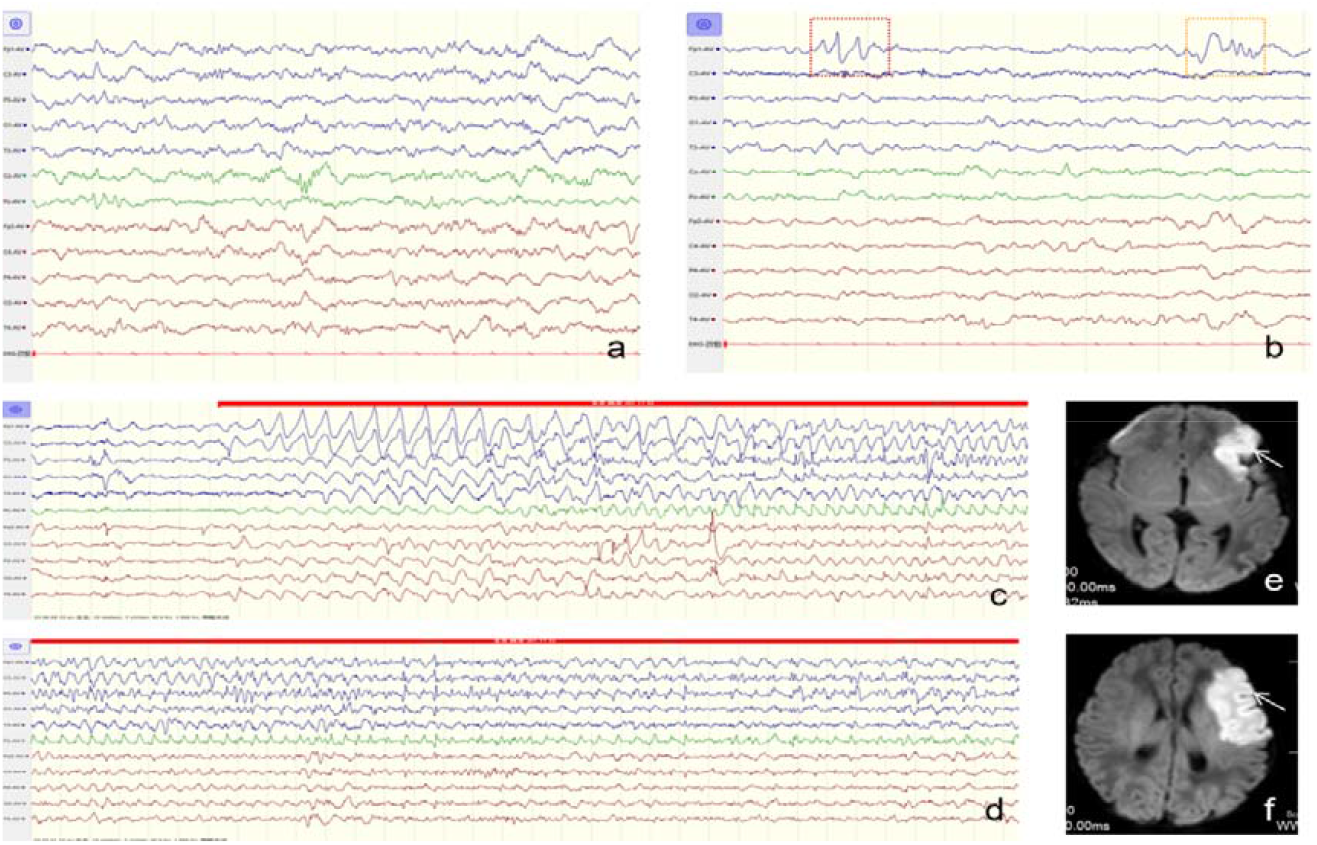
(a) Diffuse hyperactive background activity in both hemispheres, β wave activity. (b) High amplitude sharp wave and malformation wave activity in the left frontal area. (c–d) A clonic seizure, starting from the left frontal and central area, spreading throughout the left hemisphere. (e–f) Diffusion-weighted imaging hyperintensity in the left frontal area.

**Figure 2.**
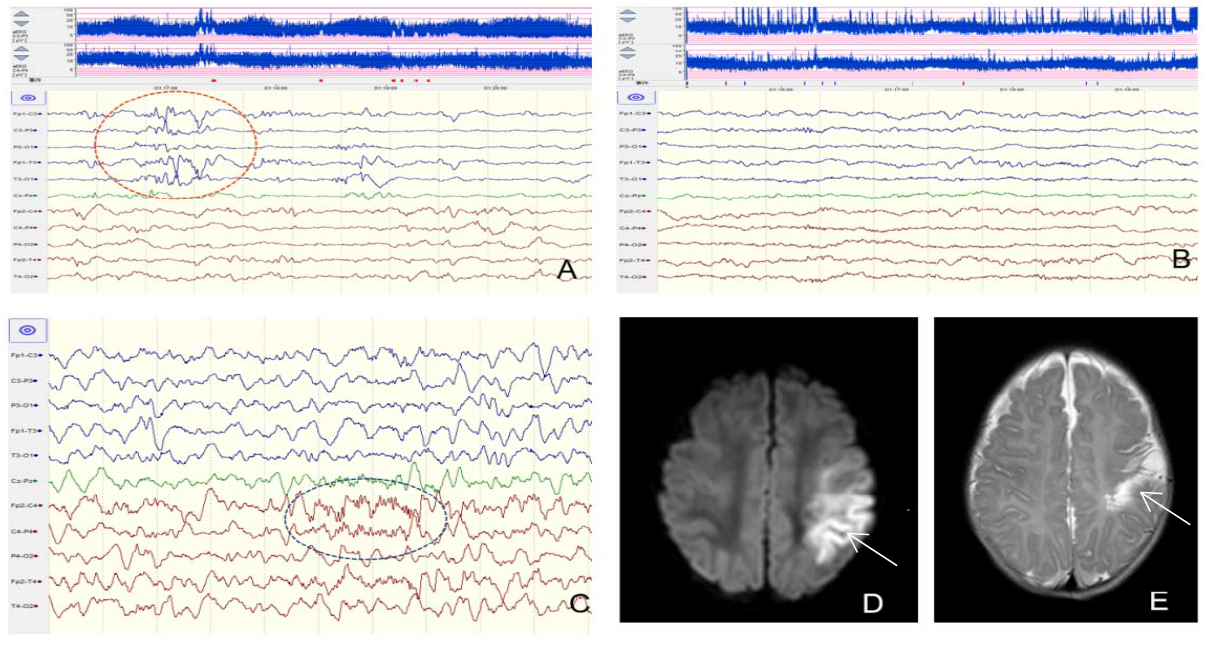
The patient was born at gestational age 40 weeks + 1, by the birth canal, with birth weight of 3,200 g. The amniotic fluid, umbilical cord, and placenta showed no abnormalities. Apgar score 1 min and 10 min, 5 min and 10 min; there was no history of asphyxia and rescue history. Seizures occurred about 13 h after birth. At 43–48 h after the onset of the disease, EEG showed multiple β wave and sharp wave activity in the left hemisphere, especially in the central and temporal regions (orange round box). Amplitude-integrated electroencephalogram (aEEG) shows a slightly lower boundary, left and right hemisphere bandwidth asymmetry, sleep–wake cycle (SWC) anomaly, and an increase in broadband period. Multiple electroclinical seizures (ECSz) (red marker) were monitored, which manifested as clonic seizures dominated by right-limb shaking. The seizures originated from the left central and central midline regions. (B) Seven days after the onset of diffuse low-medium amplitude mixed wave activity in both hemispheres, sharp wave activity decreased significantly. The left central and apical areas had low and flat wave amplitude and decreased EEG activity. Voltage and SWC were normal, with no obvious asymmetry, electrographic seizures (ESz), or ECSz. (C) One month after birth, physiological sleep spindles (round blue box) appeared in the right frontal and central areas during sleep, and no spindle appeared in the corresponding left side. (D) Seven days after birth, diffusion-weighted imaging showed high signal in left frontal, parietal, and temporal regions (indicated by arrows). (E) One month after birth, the corresponding brain regions showed encephalomalacia (arrow).

## Data Availability

The data that support the findings of this study are available from the corresponding author upon reasonable request. The corresponding author has full access to the data in this study and attests to the quality and integrity of the data set and associated analyses.

## Non-standard Abbreviations and Acronyms

(BRDs): brief rhythmic discharges
(ECSz): electroclinical seizures
(EEG): electroencephalogram
(ESz): electrographic seizures
(IBI): interburst interval
(MRI): magnetic resonance imaging
(PAIS): perinatal arterial ischemic stroke
(PDs): periodic discharges
(SWC): sleep–wake cycling
(SE): status epilepticus

## Acknowledgments

Department of Neurology, Shengjing Hospital of China Medical University, Shenyang, China (F.X.Y., W.Q). Department of Neonatal Pediatrics, Shengjing Hospital of China Medical University, Shenyang, China (W.Y.J., M.J)

